# Oropouche Virus Outbreaks in Northeast Brazil Between 2024-25 are Characterized by Sustained Transmission and Spread to Newly Affected Areas

**DOI:** 10.1101/2025.08.07.25333150

**Authors:** Elverson Soares de Melo, Sophia Maria Dantas da Silva, Gustavo Barbosa de Lima, Adalúcia da Silva, Alexandre Freitas da Silva, Verônica Gomes da Silva, Elisa de Almeida Neves Azevedo, Letícia Welter Rother, Keilla Maria Paz e Silva, Diego Arruda Falcão, Andreza Pâmela Vasconcelos, Mayara Matias de Oliveira Marques da Costa, Eduardo Augusto Duque Bezerra, Thiago Franco de Oliveira Carneiro, Erik Matthaus de Lima Paiva, Janaina Correia Oliveira, Matheus Filgueira Bezerra, Marcelo Henrique Santos Paiva, Bartolomeu Acioli-Santos, Clarice Neuenschwander Lins de Morais, Tulio de Lima Campos, Gabriel da Luz Wallau

**Affiliations:** Department of Entomology and Bioinformatics Core, Aggeu Magalhães Institute (IAM), Oswaldo Cruz Foundation (Fiocruz/PE), Recife, Brazil; Federal University of Pernambuco (UFPE), Recife, Brazil; Graduate Program in Public Health, Aggeu Magalhães Institute (IAM), Oswaldo Cruz Foundation (Fiocruz/PE), Recife, Brazil; Department of Virology, Aggeu Magalhães Institute (IAM), Oswaldo Cruz Foundation (Fiocruz/PE), Recife, Brazil; Graduate Program in Agronomy, Federal University of Santa Maria (UFSM), Santa Maria, Brazil; Central Laboratory of Public Health of Pernambuco (LACEN-PE), Recife, Brazil; Pernambuco State Department of Health, Recife, Brazil; Central Laboratory of Public Health of Paraíba (LACEN-PB), João Pessoa, Brazil; Department of Microbiology, Aggeu Magalhães Institute (IAM), Oswaldo Cruz Foundation (Fiocruz/PE), Recife, Brazil; Bioinformatics Core, Aggeu Magalhães Institute (IAM), Oswaldo Cruz Foundation (Fiocruz/PE), Recife, Brazil; Department of Arbovirology and Entomology, Bernhard Nocht Institute for Tropical Medicine, WHO Collaborating Center for Arbovirus and Hemorrhagic Fever Reference and Research, National Reference Center for Tropical Infectious Diseases, Hamburg, Germany; Federal University of Santa Maria (UFSM), Santa Maria, Brazil

**Keywords:** *Orthobunyavirus* Infections, Spatial Analysis, Phylogeography, Molecular Epidemiology

## Abstract

Oropouche virus (OROV) is a reemerging arbovirus associated with recurrent outbreaks of febrile illness, usually confined to the Amazon biome in South America. However, OROV has recently undergone an extensive expansion in Brazil, establishing local transmission in non-endemic regions, such as Northeast (NE) Brazil. The dynamics of OROV spread and a detailed analysis of sustained transmission chains and features that allow the virus to be maintained in this region are still scarce. Here, we conducted an integrated epidemiological, spatial, and molecular analysis to investigate OROV spread in Northeast (NE) Brazil from March 2024 to April 2025. Confirmed cases were analyzed in relation to ecological risk factors and geographical clustering. Additionally, we generated 65 new OROV genome sequences from the Northeast states of Pernambuco, Paraíba, and Sergipe, and inferred the spatiotemporal dynamics of the virus in the region. A total of 2,806 confirmed cases were reported in Northeast Brazil during the study period, affecting 170 municipalities across eight out of nine NE states, in highly heterogeneous incidence patterns. A notable ecological shift was observed, with OROV transmission moving from Atlantic Forest areas in 2024 to humid zones of Caatinga biome in 2025. Phylogenetic reconstruction revealed multiple independent viral introductions in Northeast in 2024, two of them into Pernambuco. The first, derived from the central region of Amazonas, became the main driver of local transmission and subsequently spread to Paraíba and Sergipe, causing outbreaks. The second, from Rio de Janeiro, remained restricted within Pernambuco. Most cases in Sergipe and Pernambuco were restricted to 2024, while in Paraiba, most cases were recorded in 2025 following a single introduction from Pernambuco. These findings indicate a spatiotemporal displacement of the OROV outbreak in these neighboring states. While several municipalities reported high OROV incidence, Jaqueira (Pernambuco) emerged as a key hub for regional viral spread, with human mobility likely playing a key role in the interstate OROV spread. Despite a shared viral origin, each state experienced a distinct epidemic dynamic, due to different ecoclimatic conditions, short-lived and fast locally spreading outbreaks.

**HIGHLIGHTS:**

- OROV spread quickly in Northeast Brazil due to multiple independent introductions.
- Pernambuco emerged as a dissemination hub, spreading OROV to neighboring states.
- OROV crossed ecological boundaries, reaching humid regions of Brazil’s Caatinga biome.

## INTRODUCTION

The *Orthobunyavirus oropoucheense* (OROV), the causative agent of Oropouche fever, is an arbovirus belonging to the family Peribunyaviridae, and primarily transmitted by the biting midge *Culicoides paraensis* ^1^. First detected in 1955 in Trinidad and Tobago during an outbreak of acute fever among forest workers, it has since become one of the leading causes of arboviral infections in Latin America ^2^. Until 2012, OROV was the second most frequent arbovirus in Brazil, surpassed only by the Dengue virus. It was later surpassed by the viruses responsible for Chikungunya and Zika fevers ^3^. Since its discovery, there have been more than 30 OROV outbreak events, mostly in South America. However, travel-associated cases have also been identified on other continents, including in Central America (e.g., Cuba) and Europe (Spain, Germany, and Italy) ^2,4^.

Oropouche fever causes a range of symptoms like high fever, headache, myalgia, and arthralgia, and less frequently, rash, retro-orbital pain, and anorexia. The similarity in clinical symptoms often complicates the differentiation between Oropouche fever and other arboviruses circulating in Brazil, such as Dengue, Chikungunya, and Zika. In rare cases, OROV can also reach the central nervous system ^5^, and there are reports of its potential link to congenital malformations, for example, microcephaly, similar to what is seen with fetal Zika virus infection ^6^. Lastly, viral RNA has already been detected in multiple fetal tissues following intrauterine death, where OROV was identified as the cause, with findings suggesting the development of symptoms similarly to those caused by Zika virus (ZIKV) infection ^7,8^.

OROV is a tri-segmented virus having the S – small, M – medium and L – large segments coding for nucleocapsid protein and a non-structural protein, glycoproteins and a non-structural protein, and the polymerase RdRp, respectively ^9^. As such, this virus evolves both by antigenic drift, accumulation of non-synonymous mutations that may alter the structure of the viral antigen, and reassortments which occur when two different viral lineages co-infect the same host, giving origin to a lineage with a new segment composition ^10^. Phylogenetic reconstruction conducted before the current outbreak suggested that the OROV genome has undergone various reassortment events over the years. The S and L segments emerged in the first half of the 20th century, while the M segment is older, dating back to the early 17th century ^11^. As of 2023, three reassortant OROV lineages were known, all containing the L and S segments from OROV and an M segment of unknown origin: the Iquitos virus (IQTV), isolated in 1999 ^12^; the Madre de Dios virus (MDDV), identified in 2007 ^13^; and the Perdões virus (PDEV), isolated in 2012 ^14^. The acquisition of a new M segment usually leads these viruses to encode a different glycoprotein, the virus’s main antigenic protein, which can reduce the immune protection conferred by prior OROV infections and change host receptor binding features of the virus ^3^.

Over the past two years (2023–2024), the Americas have experienced a marked increase in the number of OROV cases, with 313 reported in Bolivia, 259 in Peru, 38 in Colombia, 74 in Cuba and over 5000 in Brazil as of May 2024 ^15^. In Brazil, this increase of cases was first detected in the Northern region of the country, which includes the states of Amazonas, Acre, Roraima and Rondônia ^16^. A new viral lineage was found underlying this rise in infections. This lineage results from the reassortment between the M segment of viral strains previously circulating in the Northern Region and the L and S segments derived from viruses sampled in neighboring countries such as Colombia, Peru, and Ecuador. It is estimated that this new lineage reemerged around 2013– 2014 and circulated for approximately nine years without triggering major human outbreaks, until a 2022-24 wave was detected. This led to the emergence of four distinct OROV clades by early 2024, designated as AMACRO-II, AM-I, AM-II, and AM-III ^16^. Subsequently, OROV infection spread to several non-Amazonian Brazilian states, generating a national public health emergency in 2024. Phylogeographic analyses supported evidence of active transmission outside the Northern region and also indicated single or multiple introductions in a number of Brazilian states ^17^. As of July 4, 2024, Brazil had recorded 6,976 confirmed cases of Oropouche Fever, with 21.6% of cases occurring outside the Amazon region ^18^.

In the Brazilian Northeast region, a single introduction event of OROV into the state of Pernambuco was described by Gräf et al. (2024) and Ceará by Lima et al. (2024) ^17,19^, and both lineages originated directly from the state of Amazonas. Nevertheless, additional introduction events may have gone undetected due to limited sampling, notably the dynamics underlying the dissemination and maintenance of OROV across states in the Northeast region poorly understood. Moreover, the main drivers of OROV sustained transmission in previously non-endemic regions remain to be uncovered. Here, we investigated the epidemiological, spatial, and ecological dynamics of Oropouche fever across Northeast Brazil between 2024 and 2025. We characterized patterns of incidence, spatial clustering, and ecological risk factors associated with OROV transmission at the municipal level. Additionally, we sequenced OROV genomes directly from clinical samples from Northeast states from Brazil (Pernambuco, Paraíba, and Sergipe) and performed molecular and Bayesian phylogeographic analysis. Our findings uncovered a new introduction event in Pernambuco and revealed that the Pernambuco outbreak served as a key dissemination hub for the spread of the virus to neighboring states such as Paraíba and Sergipe. OROV has sustained transmission in the region over a two-year period, characterized by marked spatiotemporal displacement consistent with short-lived, rapidly spreading outbreaks, followed by cryptic transmission and subsequent dissemination to new areas, ultimately driving renewed intense outbreaks.

## METHODS

### Epidemiological data collection and spatial analysis

Data on the number of Oropouche Fever cases per municipality in the Northeast region, along with their corresponding epidemiological reporting weeks, were retrieved from the Oropouche Fever Epidemiological Dashboard maintained by the Brazilian Ministry of Health ^20^. To calculate the incidence rate per 100,000 inhabitants for each municipality, we used the 2024 municipal population estimates provided by the Brazilian Institute of Geography and Statistics (IBGE) ^21^. The analysis period spanned from March 2024 to April 2025, corresponding to the first confirmed cases in the region. This interval was selected to capture the full course of the outbreak from its emergence, allowing for a comprehensive evaluation of the incidence and geographic spread of the disease. Agricultural production data for the year 2023 were obtained from the IBGE ^22^. Spatial data, including the shapefiles of municipal and intermediary regions (group municipalities based on urban networks and regional influence), biomes, vegetation area of 2023, and land cover and use of 2024, were also obtained from the IBGE ^23^. We also extracted the most recent data about land cover and use from the MapBiomas project ^24^.

To mitigate incidence instability in municipalities with small populations, spatial empirical Bayes smoothing was applied to municipal-level incidence rates using TerraView software (version 4.4) ^25^. Geographic data integration and the creation of thematic maps showing the number of cases, crude incidence, and smoothed incidence were carried out using QGIS (version 3.40) ^26^.

Finally, we conducted spatial autocorrelation analysis using GeoDa software ^27^ (version 1.20). We applied the Empirical Bayes (EB) adjustment for Moran’s I as proposed by Assunção and Reis ^28^, implemented in GeoDa as the “Moran’s I with EB Rate” procedure. Global and local Moran’s I statistics were calculated to identify spatial patterns and clusters of high and low incidence, considering first-order neighbors (queen criterion). Statistical significance was assessed using Monte Carlo permutation tests (n = 999), with results deemed significant at a pseudo p-value <0.05.

### OROV sample collection and whole-genome sequencing

Initial detection of Oropouche Fever cases was carried out by the Central Public Health Laboratories of Pernambuco (LACEN-PE), Sergipe (LACEN-SE), and Paraíba (LACEN-PB) using a real-time reverse transcription PCR (RT-qPCR) assay targeting OROV. Serum samples from RT-qPCR–positive patients were sent to the Arbovirus Reference Service at the Aggeu Magalhães Institute (FIOCRUZ Pernambuco), where they were stored at –80 °C until viral RNA extraction. For subsequent molecular analysis, only samples with a Ct (cycle threshold) value below 30 were selected for RNA extraction and sequencing.

Viral RNA was extracted using ReliaPrep Viral Total Nucleic Acid Purification Kit (Promega), library preparation was performed with the Illumina COVIDSeq backbone, offering a cost-effective genome-wide sequencing solution. The amplification of complete OROV genomic segments was performed through the AmpliSeq method, using a set of OROV-specific primer panels, as described by Naveca et al. (2024). High-throughput sequencing was carried out on the MiSeq platform using a 150 bp paired-end approach.

This study received ethical approval from the ethics committee of Instituto Aggeu Magalhães (CAAE - 10117119.6.0000.5190).

### Genome assembly

For the reconstruction of the viral genome sequences sequenced reads were processed using the ViralFlow pipeline (version 1.2.0) ^29^, which implements a reference-guided genome assembly approach. The reference sequences used correspond to the L (Accession: OL689334.1), M (Accession: OL689333.1), and S (Accession: OL689332.1) segments from the lineage causing the outbreak in Brazil. Samples with less than 70% coverage in any of the three segments were excluded from subsequent phylogenetic and phylodynamic analyses.

### Segment-Based Phylogenetics and Recombination Filtering

To perform the evolutionary analysis, in addition to the genomes from samples collected in the states of Pernambuco, Paraíba, and Sergipe, viral genomes from samples collected in other Brazilian localities were also retrieved from public databases NCBI and GISAID (Global Data Science Initiative) ^30^ from 2022 up to January 30, 2025, particularly sequences produced by Naveca et. al. (2024), Gräf et al. (2024), Moreira et al. (2024) and Lima et al. (2025). Each segment was aligned independently using the MAFFT tool ^31^ (version 7.490), followed by manual curation to remove non-coding sequences and unaligned regions. The RDP5 ^32^ software was used to detect recombinant regions, and no regions were found.

Phylogenetic trees for each viral genome segment were initially constructed using the Maximum Likelihood method implemented in IQ-TREE ^33^ (version 2.3.6), with evolutionary model selection performed by ModelFinder ^34^ and applying 1,000 bootstrap replicates. Tree topologies were compared to identify potential phylogenetic incongruities suggestive of new segment reassortment events. As no new reassortments were found, all sequences were used in downstream analysis using concatenated segments.

### Temporal Signal examination

For the phylodynamic analysis, the three viral segments were concatenated using the SeqKit tool ^35^, resulting in a representative genome for each sample. As with the individual segments, the concatenated genomes were also aligned using MAFFT, and phylogenetic inference was performed with IQ-TREE. To assess the temporal signal and molecular clock assumption, root-to-tip regression analysis was performed using TempEst ^36^ (version 1.5.3), which calculates and visualizes the relationship between sampling time and genetic divergence.

### Phylodynamic and phylogeographical analysis

To reconstruct the spatiotemporal dynamics of OROV in Northeast Brazil, focusing in particular on municipalities in the states of Pernambuco, Paraíba and Sergipe, we constructed a discrete time-calibrated phylogenetic tree using BEAST v1.10.4 ^37^. The evolutionary model applied was GTR+G+I, as recommended by ModelFinder, and codon positions were treated as separate partitions to account for substitution rate variation across sites. An uncorrelated lognormal relaxed molecular clock was employed to allow rate variation among lineages. As a tree prior, we adopted the coalescent Bayesian Skyline model, which allows for the inference of changes in effective population size over time without relying on strict parametric assumptions regarding viral demography. We defined geographic locations as discrete traits for analysis. These included: (i) grouped cluster comprising the three states of Brazil’s Southern region; (ii) a grouped cluster comprising Acre, Rondônia, and southern municipalities of Amazonas; (iii) a set of individual states—Roraima, Rio de Janeiro, Espírito Santo, São Paulo, Amazonas, Ceará, Bahia and Sergipe; and (iv) municipalities reporting OROV cases within Pernambuco and Paraíba, which were analyzed separately due to their local importance in viral dispersion. An asymmetric discrete trait substitution model was applied, combined with a Bayesian Stochastic Search Variable Selection (BSSVS) procedure to limit the number of transition rates to only those supported by the data, thereby enhancing the interpretability of the phylogeographic diffusion patterns.

To ensure robustness in the phylogenetic inference, ten independent Markov Chain Monte Carlo (MCMC) runs were conducted using the Metropolis-Hastings algorithm, each comprising 100 million generations. After completion of all runs, the chains were combined using *LogCombiner* from the BEAST package, with the initial 10% of states discarded as burn-in. Convergence diagnostics were performed in *Tracer* v1.7.1 ^38^, by evaluating the Effective Sample Size (ESS) of each parameter. Only parameters with ESS values above 200 were considered reliable, indicating adequate mixing and convergence of the chains. The time to the most recent common ancestor (tMRCA) for clades associated with introductions into Northeast Brazil was estimated using the *TreeStat* tool from the BEAST package. The final Maximum Clade Credibility (MCC) tree was obtained with *TreeAnnotator* (BEAST package) and visualized using *FigTree* (https://github.com/rambaut/figtree/).

Finally, spatial diffusion patterns were assessed and visualized using SpreaD3 (Spatial Phylogenetic Reconstruction of Evolutionary Dynamics) ^39^, which enabled the spatiotemporal interpretation of viral movement across geographic regions.

## RESULTS

### Epidemiological Dynamics of Oropouche Fever in the Northeast of Brazil

According to the Brazilian Ministry of Health, 834 confirmed cases of Oropouche fever occurred in 2023, most of them in states of the Amazon basin in Northern Brazil (Fig. 1). By 2024, this number had increased to approximately 13,800 cases, with autochthonous cases reported in 22 Brazilian states, while by April 2025, around 11,300 cases had already been confirmed, indicating an acceleration in the spread of the virus (Fig. 1B). Within this epidemiological scenario, the Northeast region of Brazil recorded 1,517 cases of Oropouche fever in 2024 (~11% of national cases), distributed across 8 states, and 1,289 cases by May 2025, reported in 5 states (~11,4% of national cases). The sex distribution of cases in the region was approximately balanced, with 50.32% female, 49.54% male, and 0.14% with unreported sex. Regarding age distribution, most cases (62.19%) occurred among the 20 to 49 age group.

**Figure 1.**
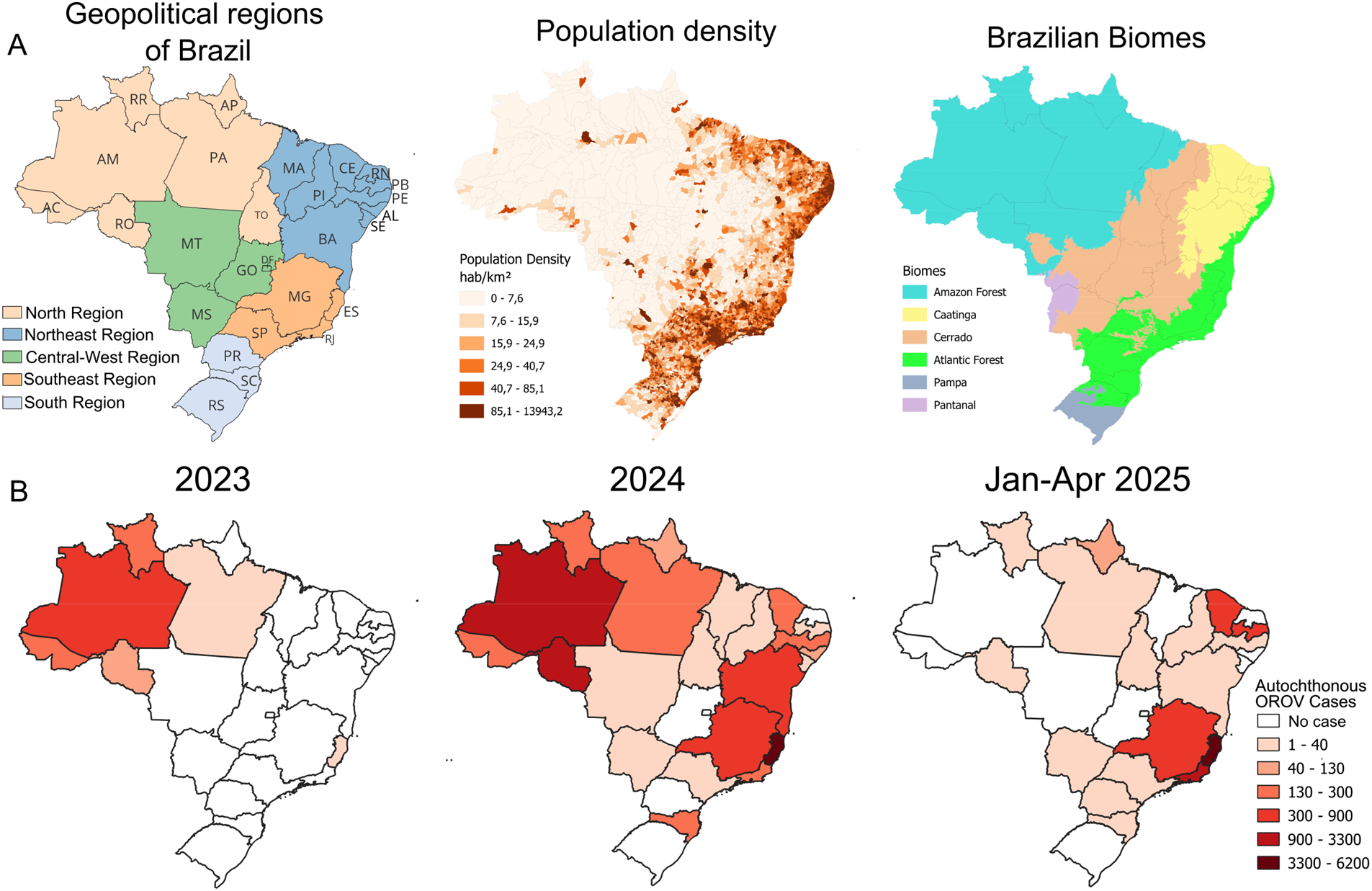
Geographic and temporal distribution of autochthonous OROV cases across Brazilian states (January 2023 to April 2025) in relation to geopolitical regions, population density and biomes. (A) The maps illustrate the current geopolitical division of Brazil, highlighting regions and states, population density distribution, and biomes across the country. (B) The maps illustrate the growing geographic spread and increase in the number of autochthonous cases over time, highlighting a notable expansion from the Amazon basin in 2023 to several states in Northeast and Southeast Brazil in 2024 and 2025. Acre (AC), Alagoas (AL), Amapá (AP), Amazonas (AM), Bahia (BA), Ceará (CE), Distrito Federal (DF), Espírito Santo (ES), Goiás (GO), Maranhão (MA), Mato Grosso (MT), Mato Grosso do Sul (MS), Minas Gerais (MG), Pará (PA), Paraíba (PB), Paraná (PR), Pernambuco (PE), Piauí (PI), Rio de Janeiro (RJ), Rio Grande do Norte (RN), Rio Grande do Sul (RS), Rondônia (RO), Roraima (RR), Santa Catarina (SC), São Paulo (SP), Sergipe (SE) e Tocantins (TO).

Autochthonous transmission in Northeast Brazil was first documented in March, during the eighth epidemiological week of 2024, with two cases reported in Maranhão (MA) and one in Bahia (BA) (Fig. 2A). In Maranhão, cases were characterized by a scattered distribution between weeks 8 and 32. However, a marked increase in the number of cases was observed in Bahia following the initial case along with a small outbreak in Piauí between the 11th and 19th weeks. In Bahia, transmission persisted until the 21st epidemiological week (late May), followed by a gradual decline (Fig. 2A). Concurrently, an increase in case numbers was observed in Pernambuco (PE), where the first autochthonous case was identified in the epidemiological week 16 (mid-April 2024). The state experienced a peak in infections during the week 26 (late June), near the end of the rainy season, with the outbreak persisting through the week 38 (September). Following the peak of cases in Pernambuco, significant increases were subsequently observed in Alagoas (AL) and Ceará (CE) beginning in early July (at the beginning of the dry season). Although local transmission had already been established in both states by mid-May, the most pronounced growth occurred during the second half of the year. In Ceará, cases continued to be reported through the end of 2024; however, a declining trend was noted starting in the epidemiological week 34 (late August), mirroring the pattern observed throughout the Northeast region. Interestingly, this steep decline coincided with the months that have the lowest average precipitation (Fig. 2B). Despite this, the first two months of the dry season still allowed sustained transmission in Sergipe (SE), between weeks 25 and 36, and contributed to the continued spread of cases in Alagoas and Ceará.

**Figure 2.**
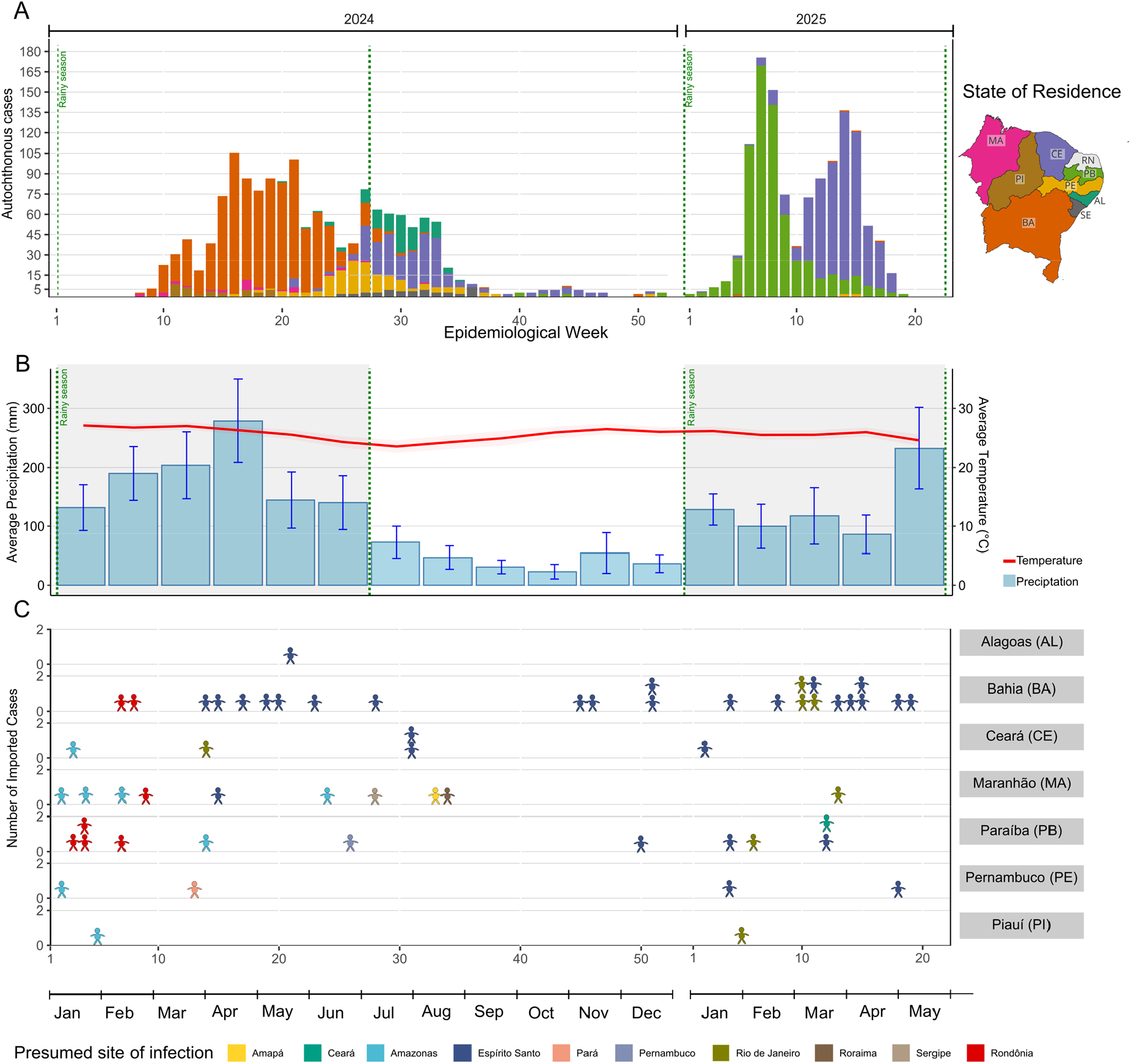
Temporal distribution of autochthonous and imported OROV cases in Northeast Brazil by epidemiological week (2024–2025). (A) Weekly distribution of autochthonous OROV cases by state of residence. The colored bars highlight how the timing and magnitude of outbreaks varied across different states from 2024 to May 2025. (B) Average monthly precipitation (blue bars) and temperature (red line) in the areas reporting OROV cases in Northeast Brazil between January 2024 and May 2025. Error bars represent 95% confidence intervals in monthly precipitation. The shaded area highlights the rainy season. (C) Weekly distribution of imported OROV cases, according to the state of residence (y-axis) and the presumed site of infection (color-coded by local of presumed infection).

Although the first autochthonous cases were officially reported in February 2024, initial cases of OROV infection among residents of Brazil’s Northeast region were documented between the epidemiological weeks 2 and 5 of 2024 (Fig. S1). Individuals from Pernambuco, Paraíba, Ceará, Maranhão, and Piauí, likely acquired the infection while traveling to the Amazonian region (**Fig. 2C**). However, it was not possible to confirm whether these cases were directly related to local transmission, due to the likely underreporting of early OROV cases at the onset of the outbreak in the Northeast region and the consequent absence of early genomic data from these patients.

The last Northeast state to report autochthonous transmission was Paraíba, with the first locally acquired case confirmed during the 40th epidemiological week of 2024 (early October).

Only six locally transmitted cases were recorded in 2024; however, a marked increase in incidence was observed beginning in the first epidemiological week of January 2025. Case counts peaked at over 165 per week by epidemiological week 7 of that year. After this peak, a reducing trend in incidence was observed (**Fig. 2A**). Concurrently with the decline in Paraíba, a new rise in cases emerged in the state of Ceará, along with additional case reports, but with comparatively lower incidence in Bahia and Pernambuco. Particularly for Ceará, in which the 2025 outbreak was much larger than in 2024, cases were again increasing in the same municipalities affected in 2024.

### Municipal Patterns of Oropouche Fever Incidence in Northeastern Brazil

In Northeast Brazil, Oropouche fever cases were reported in 170 municipalities, representing 9.84% of all municipalities in the region. The spatial distribution was heterogeneous across states, with low municipal coverage within each state (**Fig. 3A**). The lowest municipal coverage was observed in Piauí (2.77% of municipalities), while the highest occurred in Pernambuco (15.68%) (**Table 1**). Despite the higher proportion of affected municipalities in Pernambuco, the highest state-level incidence was recorded in Paraíba, with 15.54 cases per 100,000 population. In addition to these differences in municipal coverage and state-level incidence, a marked intra-state heterogeneity in municipal incidence rates was observed. In all states analyzed, municipal incidence coefficients of variation (CV) were high, indicating that transmission was concentrated in a few municipalities, while most reported low case numbers.

**Table 1.** State-level distribution, incidence rates, and spatial heterogeneity of Oropouche fever in Northeast Brazil.

| State | % of municipalities with cases<sup>1</sup> | State-level incidence (cases per 100,000 inhabitants) | Highest municipal incidence | Mean incidence<sup>2</sup> | Empirical Bayes incidence mean<sup>3</sup> | Coefficient of variation (%)<sup>4</sup> |
| --- | --- | --- | --- | --- | --- | --- |
| Maranhão | 8.75 | 0.52 | 45.6 | 9.45 | 17.83 | 344 |
| Alagoas | 13.72 | 3.73 | 131.8 | 20.16 | 28.75 | 485 |
| Bahia | 15.11 | 6.03 | 329.8 | 56.30 | 132.0 | 335 |
| Ceará | 8.15 | 8.98 | 1544.64 | 271.78 | 589.4 | 358 |
| Paraíba | 7.62 | 15.54 | 1800.82 | 192.37 | 449.4 | 473 |
| Pernambuco | 15.68 | 1.54 | 648.67 | 31.79 | 90.0 | 738 |
| Piauí | 2.67 | 0.92 | 94.1 | 41.27 | 114.86 | 342 |
| Sergipe | 9.33 | 1.44 | 225.45 | 36.71 | 123.66 | 454 |

**Legend:** <sup>1</sup> Proportion of municipalities with at least one confirmed case of the disease. <sup>2</sup> Mean of municipal incidence rates, considering only municipalities with at least one case. <sup>3</sup> Mean of municipal incidence rates smoothed using the Spatial Empirical Bayes method, considering only municipalities with at least one case. <sup>4</sup> Calculated using Empirical Bayes smoothed rates to minimize overestimation of heterogeneity due to small population sizes.

**Figure 3.**
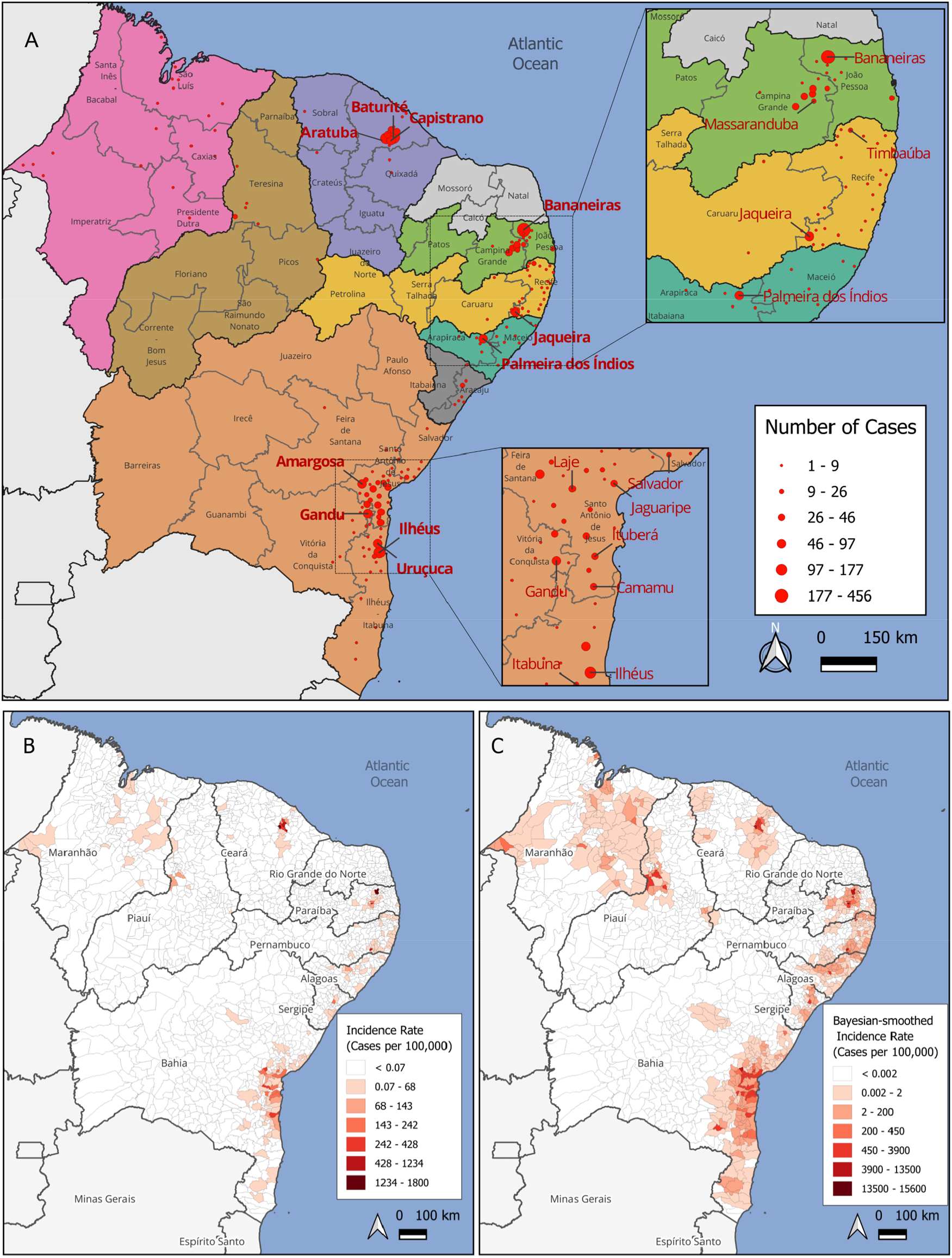
Spatial distribution and incidence rates of OROV cases in Northeast Brazil. (A) Absolute number of reported OROV cases per municipality between March 2024 and April 2025. The map is subdivided by states (color-filled shapes) and Intermediary Regions (IRs). Red circles represent the centroid of each municipality with reported cases. Labeled cities correspond to those with more than 46 cases in the main map and more than 10 cases in the inset maps. (B) Crude incidence rate of OROV cases per 100,000 inhabitants at the municipal level. (C) Bayesian-smoothed incidence rate (per 100,000 inhabitants), adjusting for population size to reduce rate instability in low-populated municipalities.

Although Maranhão was the first state to report autochthonous transmission of Oropouche fever, it had the lowest incidence in the region, with 0.52 cases per 100,000 inhabitants. Cases occurred across all its Intermediate Regions (IR), with the city of Cidelândia recording the highest number of cases (6) and the highest municipal incidence (45.6 per 100,000). In neighboring Piauí, the lowest proportion of affected municipalities (2.77%) was observed, along with the second-lowest state-level incidence. Most cases were concentrated in Amarante (16 cases), and the highest municipal incidence occurred in Jardim do Mulato (94.1 per 100,000), both located in the IR of Teresina near the Maranhão border. In Bahia, which reported the highest number of cases in 2024, the most affected municipalities were Ilhéus (138 cases), Gandu (82), and Uruçuca (73), with the latter one having the highest municipal incidence (329.87 per 100,000). Most high-incidence municipalities were located south of Salvador, within the IR of Santo Antônio de Jesus and Ilhéus-Itabuna. In Alagoas, approximately 80% of cases were reported in Palmeira dos índios, the municipality with the highest incidence, situated inland near the Pernambuco border. In Sergipe, a similar pattern was observed, with most cases concentrated in Siriri, which reported 18 cases and the highest municipal incidence in the state. In Ceará, unlike Alagoas and Sergipe, cases were more geographically dispersed among neighboring municipalities, particularly in the northern part of the state. The municipalities of Aratuba, Baturité, Capistrano, Pacoti, and Mulungu recorded the highest case counts and incidences (**Fig. 3B**).

In Pernambuco, although municipal-level Oropouche fever coverage was the highest among states in the region, transmission exhibited substantial local heterogeneity. In 2024, autochthonous cases were reported in 28 municipalities, the majority of which were located in the eastern part of the state, within the Recife Intermediate Region (**Fig. 3A**). Jaqueira (68 cases; 648.6 per 100,000 inhabitants) and Timbaúba (12 cases; 25.2 per 100,000 inhabitants) together accounted for more than half of the total cases reported. Additionally, 45% of the affected municipalities recorded only a single confirmed case, reflecting the marked spatial disparity in case distribution across the state. In 2025, Goiana, a municipality located in the northern part of Pernambuco near the border with Paraíba, was the only location with confirmed local transmission up to May, suggesting limited but persistent virus circulation into the following year. Paraíba, the last state in Northeastern Brazil to report autochthonous transmission of Oropouche fever, experienced a sharp increase in cases during early 2025. In 2024, only six locally acquired cases were reported, occurring in three municipalities: Alagoa Nova (3 cases), Campina Grande (2 cases), and João Pessoa (1 case). By May 2025, however, the number of confirmed cases had grown to 664, with 17 municipalities reporting transmission, mainly in interior regions of the state. The highest burden occurred in Bananeiras, which reported both the largest number of cases and the highest incidence rate in the Northeast (1,800.8 cases per 100,000 inhabitants). Several other municipalities also reported high incidence rates compared to the regional average, including Matinhas (35 cases; 739.1 per 100,000), Alagoa Nova (39 cases; 179.5 per 100,000), Massaranduba (18 cases; 122.7 per 100,000), Lagoa Seca (27 cases; 92.9 per 100,000), Alagoa Grande (18 cases; 67.2 per 100,000), and Campina Grande (32 cases; 7.25 per 100,000).

Spatial empirical Bayes smoothing revealed underlying spatial risk patterns (**Fig. 3C**) that were not apparent in crude incidence maps, particularly in small municipalities with zero or very few reported cases. In such areas, the absence of reported cases is unlikely to reflect a true absence of risk and more likely results from underdetection due to limited surveillance capacity and statistical instability inherent to small-area data. High-incidence areas remained prominent; however, spatial smoothing revealed that most municipalities near the coastal areas of Bahia, Sergipe, Alagoas, and Pernambuco present some degree of risk for Oropouche Fever. Additional risk was highlighted in municipalities within the Campina Grande IR (Paraíba) and along the Maranhão–Piauí border, where small populations had previously masked the underlying risk. This spatial pattern suggests possible under-detection of transmission in adjacent regions where outbreaks have been detected. The names of municipalities with smoothed incidence higher than 1000 are highlighted in **Fig. S2**. We also conducted a correlation analysis between population size and the smoothed incidence rate across municipalities in the Northeast region of Brazil, which revealed a moderate negative association in the log-transformed space (r = −0.36; p < 2.2×10^−16^), indicating that more populous municipalities tend to exhibit proportionally lower incidence rates **(Fig. S3)**.

We used spatial autocorrelation methods to identify whether municipalities with similar incidence were geographically clustered. A weak but statistically significant spatial autocorrelation was detected (Moran’s I = 0.158; pseudo p-value = 0.002), suggesting a weak tendency for municipalities with similar incidence rates to be geographically clustered. The local spatial autocorrelation analysis (LISA) further identified four clusters of high disease incidence **(Fig. S4)**, located in southern Bahia (25 municipalities), Ceará (9 municipalities within the Fortaleza Health Region), Paraíba (8 municipalities within the Campina Grande Health Region), and the southern forest zone of Pernambuco (2 municipalities). In addition to these clusters, 27 municipalities were classified as Low-High spatial outliers, suggesting transitional areas with potential for transmission expansion or regions with possible case underreporting. These municipalities were mainly concentrated in five geographic areas: northern Ceará (Caridade, Canindé, Itapiúna, Aracoiaba, Ibaretama, and Acarape), southern Bahia (Nilo Peçanha, Brejões, Jiquiriçá, Varzedo, Santa Terezinha, Jaguaripe, and Buerarema), southern Pernambuco (Lagoa dos Gatos and São Benedito do Sul), Paraíba (Esperança, São Sebastião de Lagoa de Roça, Serra Redonda, Pirituba, Belém, Dona Inês, and Tacima), and Alagoas (Mar Vermelho and Belém, both neighboring Palmeiras dos índios). Nevertheless, most municipalities in the Northeast region exhibited low incidence rates and no significant spatial clustering, suggesting a limited geographic spread of the disease.

### Ecological and Climatic Determinants of OROV Case Distribution

Oropouche fever cases occurred across municipalities located within all four major biomes present in Northeastern Brazil: the Amazon Forest, Cerrado, Caatinga, and the Atlantic Forest (**Fig. 4A**). However, the distribution of cases was highly irregular. In 2024, the cases were predominantly reported in municipalities within the Atlantic Forest, following a trend observed in Brazil ^17^. However, a notable shift was observed in 2025, when approximately 98% of all confirmed cases in the Northeast region were located within the Caatinga biome. This shift represents a substantial change in the ecological profile of Oropouche fever transmission, which, prior to 2023, had been largely confined to the Amazon Rainforest biome. Moreover, most OROV cases and all four spatial autocorrelation clusters with a large number of cases are restricted to per-humid, humid, and sub-humid climate zones. Therefore, even when the cases occur within the Caatinga biome, high-risk areas remain restricted to more humid climate zones and do not extend into the semi-arid region. Additionally, no cases were reported in areas experiencing more than nine dry months per year (**Fig. S5 A**). For reference, all climatic zones and precipitation regimes across Brazil are illustrated in **Fig. S6**.

**Figure 4.**
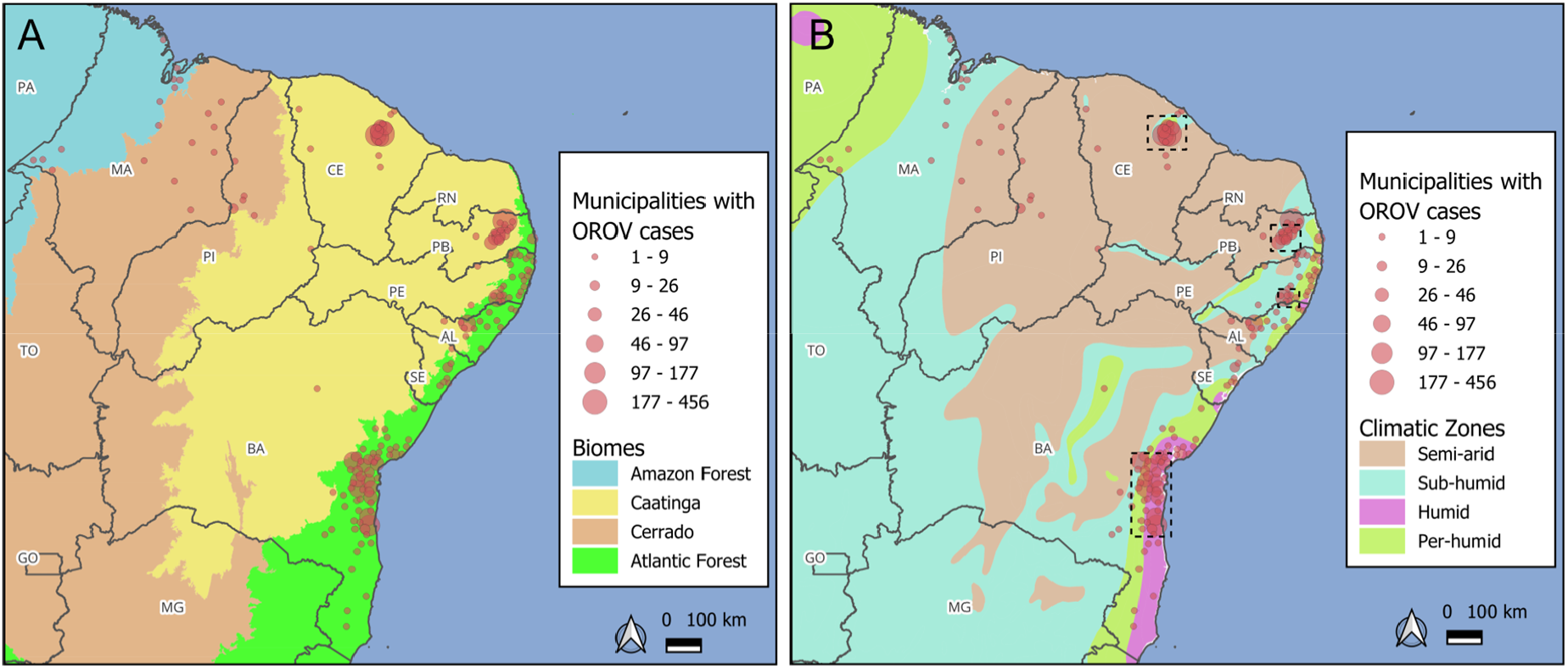
Distribution of OROV cases by biomes (A) and climatic zones (B) in Northeast Brazil. The maps show only cases reported within the Northeast region. Dashed-line rectangles in panel B indicate the four spatial autocorrelation clusters with a high number of cases, as identified by local Moran’s I analysis.

OROV cases were also reported across municipalities with diverse land use and land cover patterns, ranging from areas with extensive forest vegetation, through regions classified as mosaics of occupations in forest areas, agricultural zones, and even artificial (urban) areas **(Fig. S5 B)**. In Pernambuco, for example, approximately 35% of cases had a probable site of infection in urban areas. No preferential vegetation pattern was observed among the municipalities with the highest OROV incidence in the Northeast region **(Fig. S7)**. While some cities, such as Jaqueira (PE) and Amargosa (BA), are predominantly covered by agricultural and livestock lands, others, like Aratuba (CE) and Baturité (CE), are mainly covered by primary vegetation. Conversely, cities like Gandu (BA) and Uruçuca (BA) are largely covered by secondary vegetation, whereas cities like Capistrano and Bananeiras are covered by a mix of savanna, submontane ombrophilous forest and agricultural lands. Among five high-incidence municipalities dominated by agricultural land, each from a different state, the most cultivated crops were banana in Bananeiras (PB, 44% of planted area), sugarcane in Jaqueira (PE, 69%), corn in Capistrano (CE, 58%), cocoa in Amargosa (BA, 43%), and both corn and beans in Palmeira dos índios (AL, 32% each).

### Genomic Sampling and Phylogenetic Characterization of OROV from Pernambuco, Paraíba, and Sergipe

To improve understanding of the Oropouche fever epidemic in Northeast Brazil, we generated 65 nearly-complete OROV genomes from samples collected between March 2024 and April 2025 from three Northeastern states. This dataset included samples from Pernambuco (48 cases from 10 municipalities), Paraíba (17 cases from 8 municipalities), and Sergipe (1 case from 1 municipality). We then combined these genomes with previously published sequences from municipalities in Ceará ^19^ and Bahia ^18^, covering five of the eight Northeastern states that reported OROV cases during this period. Finally, we incorporated genomes from OROV cases reported in ten other Brazilian states in the year of 2024, representing different regions of the country.

Genotypic analysis confirmed that all 65 sequenced OROV genomes from the present study belonged to the M_1_L_2_S_2_ lineage, which spread across multiple regions of Brazil in 2024. As no evidence of segment reassortment was detected among the newly generated genomes, we concatenated the three viral segments (L, M, and S) from each sample for temporal signal assessment. A positive correlation (R^2^ = 0.81, p < 2.2 × 10^−16^) between sampling time and genetic divergence of the concatenated sequences was observed (**Fig. S8**).

Bayesian phylogenetic analysis revealed multiple independent introduction events of OROV lineages associated with outbreaks in Northeast Brazil, as well as inter-state viral dissemination within the region. We identified two distinct introductions into the state of Pernambuco (PE). The first, designated the **PE-I clade**, was directly derived from municipalities in the central region of Amazonas state (AM) (posterior probability [PP] = 100%) and was specifically linked to the **AM-I clade** (**Fig. 5A**). The second introduction, referred to as the **PE-II clade**, originated in Rio de Janeiro (RJ) state (PP = 99%), following the prior establishment of the **AMACRO-II clade** in that region. Although Lima et al. (2025) ^19^ reported that the introduction of OROV into Ceará state also originated from the state of Amazonas, and despite the limited shared border between Pernambuco and Ceará, border, our Bayesian phylogenetic analysis indicates that the strains circulating in both states belongs to the same AM-I clade but represent two independent introduction and dissemination events into Northeast Brazil, with no interstate transmission between them (Fig. 5A). Furthermore, our analysis shows that the introduction of the virus into Bahia, although derived from the **AMACRO-II clade**, represents a separate introduction event, distinct from the introduction event that gave rise to the **PE-II clade** (**Fig. 5A**).

**Figure 5.**
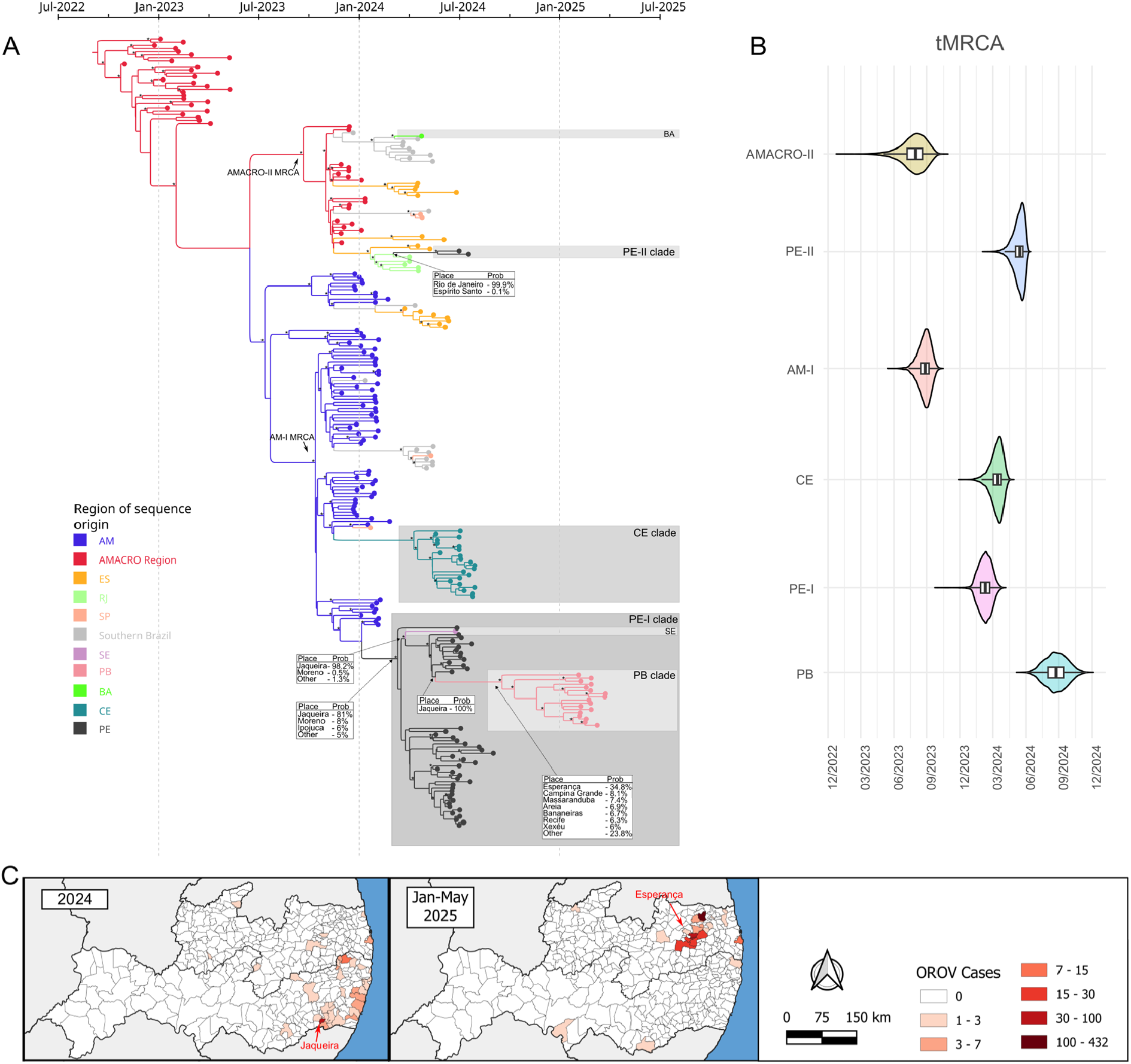
Phylogenetic and temporal dynamics of OROV clades in Northeast Brazil (2024–2025). **(A)** Time-scaled Bayesian phylogeny of OROV sequences from Brazil. Branches are color-coded by region of origin. AM = central/northern Amazonas; RO = Rondônia; AC = Acre; ES = Espírito Santo; RJ = Rio de Janeiro; SP = São Paulo; SE = Sergipe; PE = Pernambuco; PB = Paraíba; BA = Bahia; CE = Ceará; PR = Paraná; SC = Santa Catarina; RS = Rio Grande do Sul. The AMACRO Region includes AC, RO, and southern Amazonas. Southern Brazil includes PR, SC, and RS. Nodes with posterior probabilities >0.9 are marked with an asterisk (*). Northeast Brazil samples are highlighted in gray-scale squares. Tables pointing to branches represent the probabilities of ancestor origin place (**B)** Estimated times to the most recent common ancestors (tMRCAs) for key regional clades. Violin plots show 95% highest posterior density (HPD) intervals. **(C)** Geographic distribution of OROV cases (imported and autochthonous) in Pernambuco and Paraíba. The left panel shows 2024 cases, and the right panel, cases from January to April 2025. The municipalities of Jaqueira and Esperança are highlighted as initial points of viral dissemination in each period.

The estimated time to the most recent common ancestor (tMRCA) indicated that the **PE-I clade** likely emerged around February 2024 (95% highest posterior density [HPD]: December 2023–March 2024), preceding the emergence of the **CE clade**, estimated around March 2024 (95% HPD: January–May 2024), and the **PE-II clade**, which was dated to approximately May 2024 (95% HPD: March–June 2024) (**Fig. 5B**). These estimates align with the temporal pattern of case detection shown in **Fig. 2A**, suggesting that the virus circulated undetected for approximately two months before the onset of the recognized outbreak.

The PE-I clade exhibited the largest expansion within Pernambuco state and subsequently spread to neighboring states (**Fig. 5A**). The first dispersal event was from the municipality of Jaqueira to Sergipe state (PP = 98%). This was followed by a separate introduction into Paraíba state, originating from Jaqueira and possibly reaching the municipality of Esperança (PP = 34.8%). Notably, all genomes sampled from municipalities with autochthonous cases in Paraíba clustered within a single, well-supported clade (PP > 0.9), suggesting that the recent outbreak in Paraíba was primarily driven by transmission of the PE-I clade. The phylogenetic model closely reflects the observed epidemiological patterns of OROV case distribution across both states (**Fig. 5C**). Taken together, these findings point to Jaqueira as the primary hotspot of OROV transmission in Pernambuco and the likely epicenter of regional viral spread.

To illustrate the spatio-temporal progression of OROV spread across Brazil, with particular focus on the Northeast region, we reconstructed a simplified phylogeographic model highlighting the lineages that gave rise to the outbreaks in Northeast Brazil (**Fig. 6A**). Within Pernambuco, phylogeographic analyses revealed a sequential intra-state spread, primarily driven by the PE-I clade, originating in the Zona da Mata Sul region (part of the Atlantic Forest biome), specifically from the municipality of Jaqueira, which acted as the epicenter for this lineage within the state (**Fig. 6B**). The first inferred transmission from Jaqueira reached nearby coastal municipalities, including Moreno, Ipojuca, Cabo de Santo Agostinho, and Recife. During their establishment in the southern region of the state, viruses from the PE-I Clade caused fetal deaths as reported by Medeiros et al. (2025)^7^. From the coast, the virus continued its expansion toward the Zona da Mata Norte, affecting municipalities such as Aliança, São Vicente Férrer, and Timbaúba. This last city also received a second introduction of the PE-II clade from RJ. In parallel with these movements, OROV also spread from Jaqueira to other neighboring municipalities and crossed state borders to reach Sergipe—the first state within the region where cases resulting from local (intra-regional) transmission were detected. All these movements occurred between May and September 2024, reflecting a period of intense viral movement and geographic expansion.

**Figure 6.**
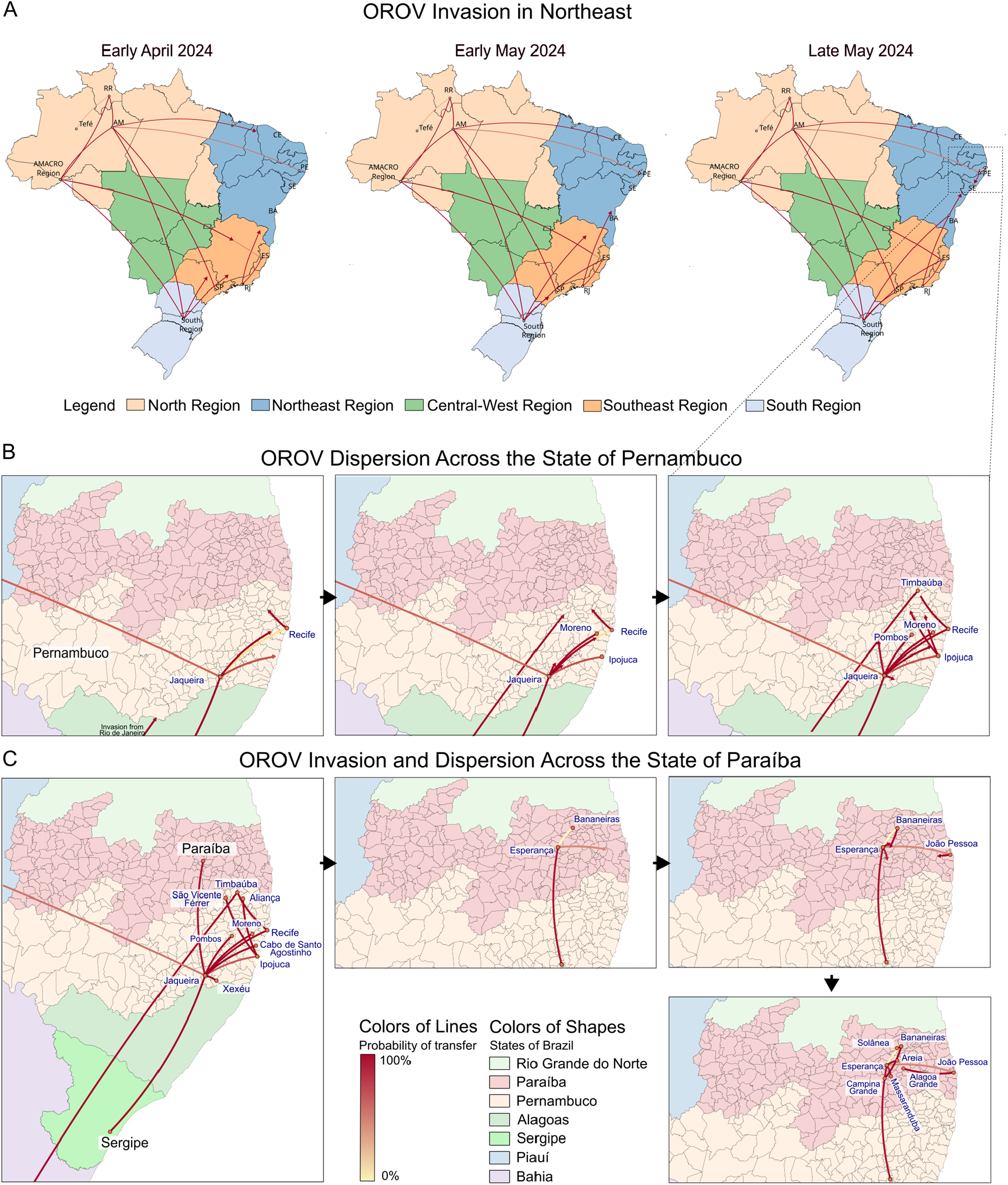
Spatial and temporal dynamics of Oropouche virus (OROV) spread across Northeast Brazil. (A) Spatiotemporal reconstruction of OROV introductions from other Brazilian regions into Northeast Brazil during April and May 2024. Maps show inferred viral movements over time, with arrows colored according to the posterior probability of each transition. Brazilian macro-regions are color-coded. (B) Inferred dispersal routes among municipalities within Pernambuco state during 2024. States in Northeast Brazil are color-coded. (C) OROV invasion in Sergipe followed by subsequent introduction and spread within Paraíba state during late 2024 and early 2025.

Jaqueira also served as a key source of viral spread into Paraíba state, with the first introduction reaching Esperança, according to our phylogeographic model (**Fig. 6C**). From there, the virus spread to nearby municipalities, including Bananeiras, Areia, Massaranduba, and the state capital, João Pessoa. Bananeiras emerged as a major outbreak hotspot, concentrating a huge number of cases and likely driving further transmission to neighboring areas. These findings illustrate how rapidly OROV expanded and formed new transmission networks in regions where it had not circulated before.

## DISCUSSION

This study highlights the rapid expansion of Oropouche virus across northeastern Brazil, a region that, until recently, had been largely unaffected by this pathogen. Between March 2024 and April 2025, both imported and local cases were detected, with sustained local outbreaks established in eight of the nine states, indicating the emergence of new potential endemic areas. The abrupt increase in case numbers, characterized by successive epidemic peaks within a short timeframe, highlights the virus’ capacity to adapt to diverse epidemiological and ecological contexts. Although most cases across Northeastern Brazil are concentrated in the rainy season, the average precipitation levels are only about 50% of the rainfall observed in the state of Amazonas during the onset of the current OROV epidemic, between 2023 and 2024 ^16^. This expansion has probably been driven by interstate population mobility and the presence of competent vectors in previously unaffected areas.

These patterns suggest an ongoing ecological diversification process, as OROV expanded into biomes and regions previously considered unsuitable for sustained transmission. Historically endemic to the Amazon region ^40^ and later detected in Atlantic Forest areas in 2024 ^17^, OROV began to show significant transmission in the Caatinga biome by 2025. The spatial spread of the currently circulating viral lineage, from the Amazon up to 2023, into the Atlantic Forest in 2024, and reaching the Caatinga in 2025, was supported by phylogeographic analyses, which revealed multiple introduction pathways from both Amazon Basin and Atlantic Forest regions. The Caatinga, a biome unique to Brazil and variably classified as a steppe savanna or dry tropical forest, is characterized by a predominantly semi-arid climate and pronounced intra-annual variability in precipitation ^41^. This ecological profile contrasts sharply with the humid tropical environments of the Amazon, which span northern Brazil and neighboring countries such as Peru, Colombia, and Venezuela, nations that also reported OROV cases in 2025 ^42^. However, Caatinga exhibits high ecoclimatic heterogeneity, and OROV cases were not uniformly distributed; instead, cases were concentrated in more humid subregions featuring patches of ombrophilous forest or arboreal or forested savanna, and areas with shorter dry seasons.

Humid and semi-humid environments favor the presence of *Culicoides paraensis*, a species that reaches high densities during the rainy season ^43^. Although widely distributed across the Americas, from the United States to Uruguay ^44^, in northeastern Brazil, this vector has been recorded only in Bahia, Ceará, Maranhão, and Pernambuco ^45^. This contrasts with recent outbreaks in states such as Piauí, Paraíba, Alagoas, and Sergipe, where no prior entomological records of the vector exist, highlighting critical gaps in entomological surveillance, as well as the possible involvement of alternative vectors, a hypothesis also raised in Venezuela ^46^. While no other *Culicoides* species have been confirmed as OROV vectors, over 150 species of the genus are present in Brazil ^45^. A study in Ceará ^47^ reported high species diversity near outbreak areas, including several anthropophilic taxa. Similar patterns were observed in municipalities bordering affected regions in Maranhão ^48^, with species that persist even during the dry season. Mosquitoes such as *Aedes serratus* have shown vector competence for other OROV lineages, though limited to sylvatic cycles ^49^. In contrast, classical urban arbovirus vectors like *Aedes aegypti* and *Culex quinquefasciatus* have demonstrated refractoriness to the currently circulating lineage under laboratory conditions ^50^, limiting their role in viral spread. Considering these findings, enhanced entomological and molecular surveillance is urgently needed in states with confirmed OROV cases but no records of *C. paraenesis*. This is essential to address knowledge gaps and identify potential alternative vectors.

Previous records indicate that the Northeast region of Brazil had already been occasionally affected by Oropouche fever. Between 1987 and 1988, a localized outbreak occurred in Porto Franco (Maranhão), within the Cerrado biome ^51^. In 2016–2017, five autochthonous cases were reported in Salvador (Bahia), but the origin of the virus remained unclear due to the absence of phylogeographic analyses and limitations in surveillance, including the lack of detection of imported cases ^52^. Although underreporting remains a significant challenge, particularly outside the Amazon region, the recent spread of OROV in Northeast cannot be attributed to the expansion of testing following the viral emergence in 2023. Genomic sequencing performed on samples from three states, Pernambuco, Paraíba, and Sergipe, along with genomes from Ceará ^19^, revealed multiple independent viral introductions in 2024. These events were associated with long-distance human migrations, primarily from Amazonas and Rio de Janeiro, both of which had active transmission during that period. Following the introduction, the virus spread locally, likely driven by routine intermunicipal movement related to work, education, and healthcare, and eventually reached neighboring states, as observed in transmission flows between Pernambuco, Sergipe, and Paraíba. The genetic sequences revealed state-specific clustering and no evidence of cross-state reintroductions, suggesting that viral circulation remained predominantly intra-state after the initial introductions. Subsequently, the disease has been established and has become more prevalent in smaller municipalities across the Northeast, particularly those located farther from major urban centers. This pattern may be associated with the presence of forested or tree-covered areas, as observed during the initial phases of OROV expansion beyond the Amazon biome in 2024 ^17^.

An average interval of approximately two months was observed between the estimated date of viral introduction in a given locality (tMRCA) and the subsequent surge in case numbers, a pattern similar to that described in other OROV introductions in Brazil ^17^. This interval may reflect the time required for infected individuals to return to their home states after traveling and initiate local transmission via vectors, combined with the viral replication period in insects and the human incubation period, which ranges from 1 to 15 days ^53^. Additionally, recent data suggest the presence of replication-competent virus in the blood of patients for up to two months following symptom onset ^54^, potentially extending the period of infectivity to vector species, a hypothesis that remains to be rigorously validated through experimental studies. The OROV tMRCA for Paraíba dates back to August/September 2024, and although few sporadic cases were recorded in the last three months of this year, the rapid growth in cases detected occurred only about 4 months later, which suggests that OROV circulated silently in Paraíba in the 2024 dry season after the wave of cases in Pernambuco had dissipated. Early testing and timely genomic sequencing are therefore critical to identify initial cases in a locality, facilitating not only the implementation of containment measures during this critical period but also the identification of viral introduction routes to inform targeted mitigation strategies.

The findings of this study highlight the urgent need to strengthen surveillance and response strategies, considering the expansion of Oropouche virus into regions previously considered non-endemic, but which already face challenges caused by other arboviruses such as DENV, ZIKV and CHIKV. The spatial smoothing approach applied in this analysis may help mitigate the effects of underreporting, yielding more robust estimates of disease risk. The results identify priority areas for syndromic surveillance of acute febrile illness, including nearly the entire coastal regions of Bahia and Pernambuco, as well as the states of Alagoas and Sergipe, in addition to specific clusters located in the interior of Paraíba and Ceará. Mapping areas at risk can guide targeted investment in the continuous training of healthcare professionals for the recognition and clinical management of suspected OROV cases, while also supporting the expansion of access to decentralized molecular testing. Strengthening local diagnostic capacity, including improvements in sample storage and RNA extraction procedures, can minimize viral RNA degradation, improve specimen quality for genomic sequencing, and reduce underreporting. Investment in rapid detection systems is particularly critical given the potential for viral reemergence in municipalities or regions that did not experience major outbreaks but possess climatic and ecological conditions favorable to *Culicoides* spp. proliferation. In such settings, the low recorded incidence is likely insufficient to establish herd immunity. Lastly, randomized seroepidemiological surveys in affected municipalities are recommended to more accurately estimate prior population-level exposure and, consequently, to assess the risk of future outbreaks based on antibody prevalence.

This study has several limitations. Given that Oropouche fever has historically been underreported in many parts of Brazil and presents symptoms that resemble those of other arboviral infections, it is likely that numerous cases were not identified by local health systems, particularly in small municipalities with limited diagnostic capacity and healthcare infrastructure. To partially address this bias, we applied a spatial smoothing approach, which provided more robust estimates of disease risk across different locations in the Northeast region. However, it is likely that several patients were asymptomatic, and municipal hubs may also be underestimated. This limitation may have led to an underestimation of priority areas for future surveillance. Additionally, it was not possible to obtain viral genomes from all affected municipalities in the states of Pernambuco, Paraíba, and Sergipe. The phylogeographic model also relied on genome sequences previously generated by other research groups for Bahia and Ceará, and did not include data from Maranhão and Piauí. This may have led to an underrepresentation of viral introduction routes across the broader Northeast region.

## CONCLUSION

This study provides robust evidence of the rapid and sustained transmission of Oropouche virus (OROV) across Northeast Brazil, driven by multiple independent introductions and local amplification networks. By integrating high-resolution genomic data with advanced spatial analyses, we identified critical dispersal hubs, such as the municipality of Jaqueira in Pernambuco, that played a central role in facilitating widespread regional dissemination. The combined analytical approach demonstrates how population mobility, favorable ecological niches, and immunologically naive populations synergistically contributed to the accelerated expansion of OROV into newly emergent endemic zones. These findings underscore the urgent need to enhance decentralized molecular diagnostic capacity, strengthen entomological and syndromic surveillance systems, and invest in healthcare workforce training across newly affected regions. The continued integration of genomic epidemiology and spatial analysis, as exemplified in this study, offers a powerful framework for anticipating and mitigating future outbreaks through more targeted and effective public health interventions.

## Data Availability

All data produced in the present work are contained in the manuscript.

## ACKNOWLEDGMENTS

We thank the FIOCRUZ Genomic Surveillance Network for their continued support, the Brazilian Ministry of Health for providing access to case data through its official portal, the public health and medical teams across Brazil for their response to the Oropouche outbreak, and the Bioinformatics Center of the Aggeu Magalhães Institute for their support.

## DATA AVAILABILITY

The sequences generated in this study have been deposited in the GISAID database and are available under the following EPI SET IDs: EPI_ISL_19542793 to EPI_ISL_19542820, EPI_ISL_19556950 to EPI_ISL_19556964, EPI_ISL_19681726, EPI_ISL_19681727, EPI_ISL_19492156, EPI_ISL_19492157, EPI_ISL_19850128, EPI_ISL_19850126, EPI_ISL_19850137, EPI_ISL_19850149, EPI_ISL_19850146, EPI_ISL_19850158, EPI_ISL_19850155, EPI_ISL_19850134, EPI_ISL_19850167, EPI_ISL_19850131, EPI_ISL_19850164, EPI_ISL_19850143, EPI_ISL_19850176, EPI_ISL_19850140, EPI_ISL_19850173, EPI_ISL_19850152, EPI_ISL_19850161, EPI_ISL_19850170.

