## Supplementary Material for "Oropouche Virus Outbreaks in Northeast Brazil Between 2024-25 are Characterized by Sustained Transmission and Spread to Newly Affected Areas" (from another version of this preprint: version 1, 2025-08-08)

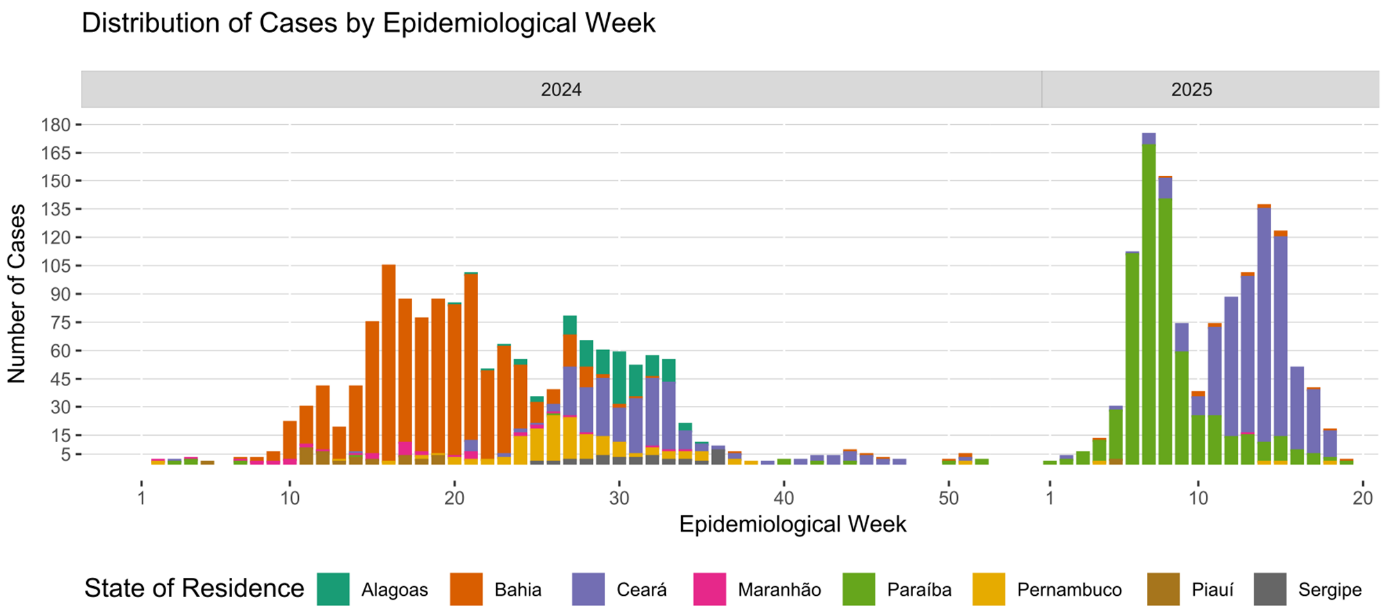


**Figure S1 -** Weekly distribution of autochthonous and imported OROV cases by state of residence, between 2024 and 2025.

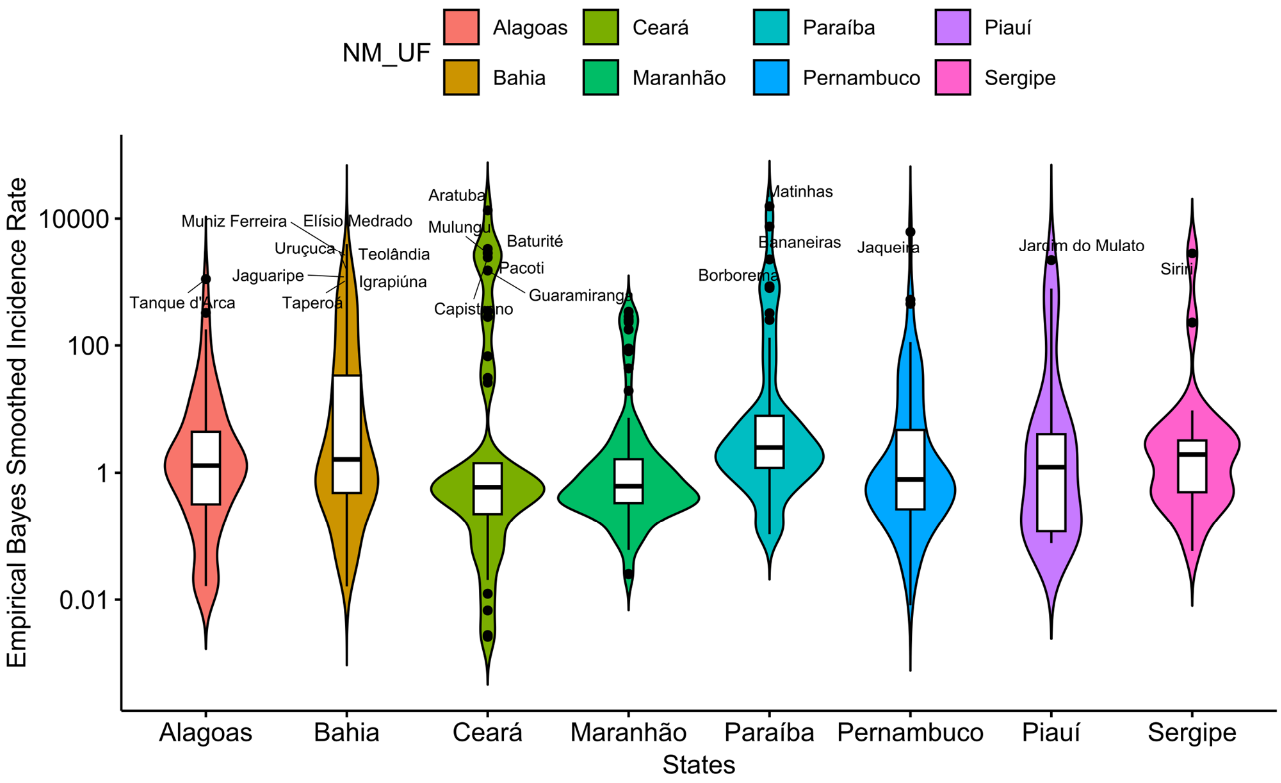


**Figure S2 -** Smoothed incidence rates of Oropouche virus by state in Northeast Brazil. Each violin plot represents the distribution across municipalities whose smoothed incidence is greater than zero. The names of municipalities with smoothed incidence higher than 1000 are highlighted.

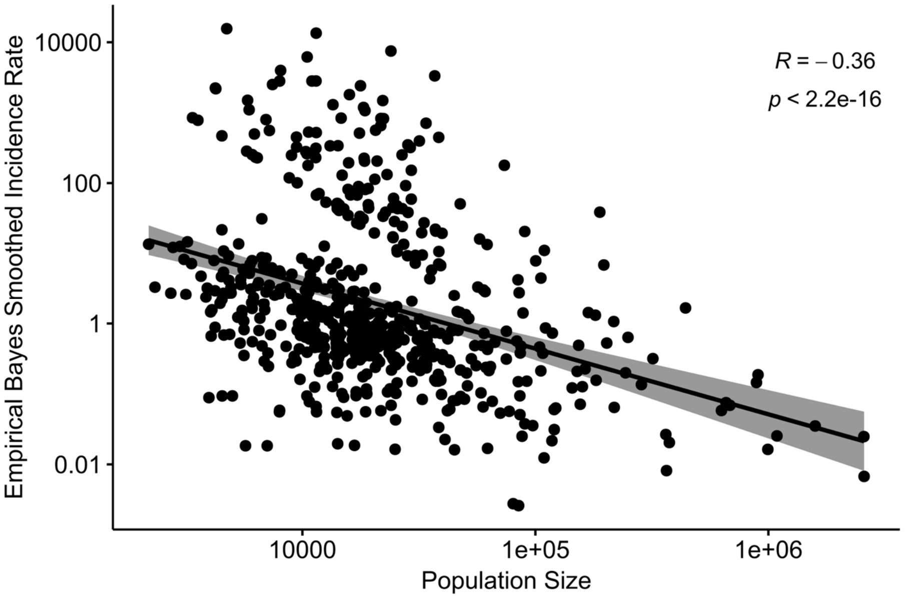


**Figure S3 -** Correlation analysis between population size and the Empirical Bayes smoothed incidence rate across municipalities in the Northeast region of Brazil. While the association is approximately linear on the logarithmic scale, it reflects a nonlinear inverse relationship in the original data distribution.

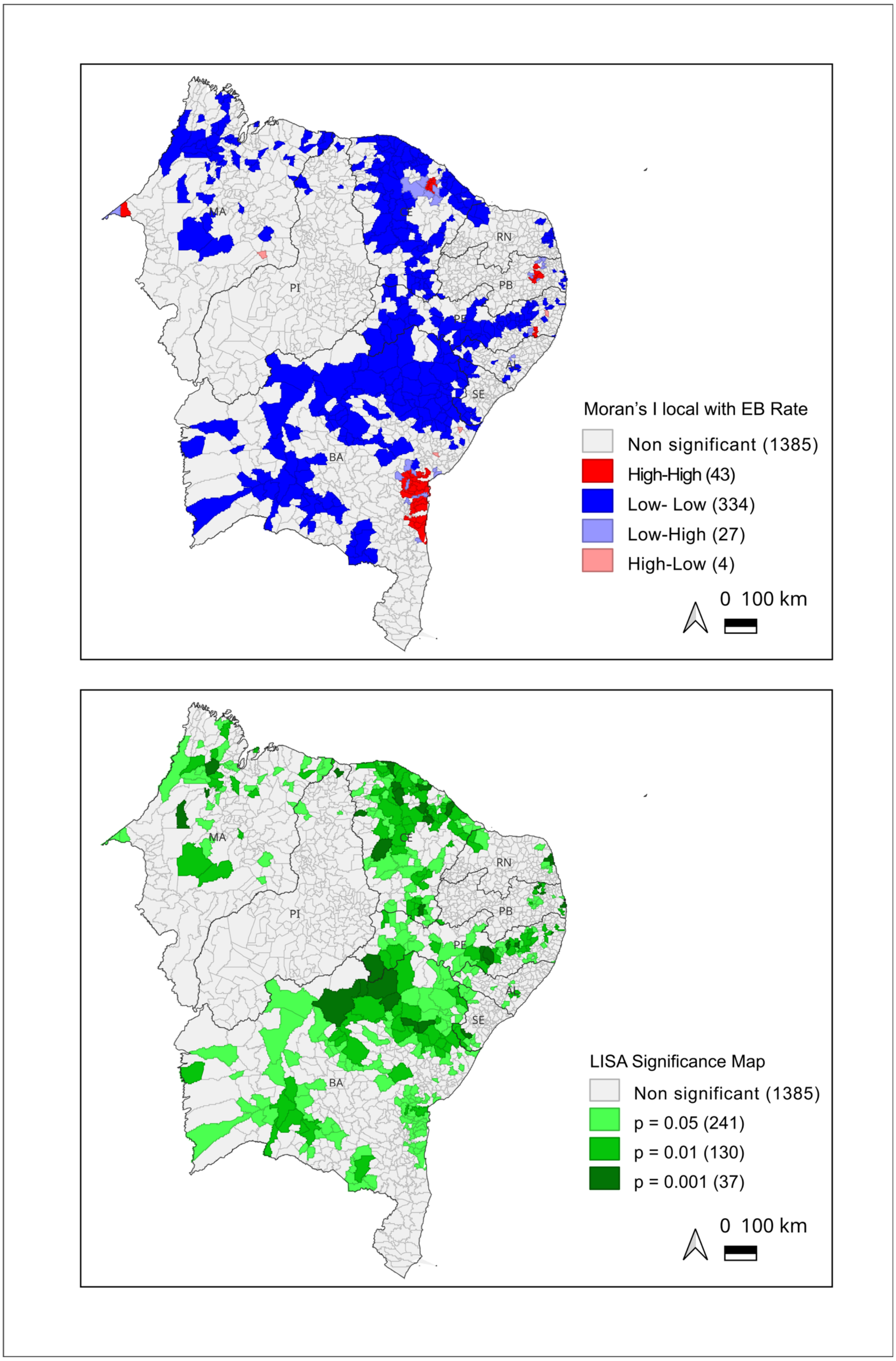


**Figure S4** - Local Indicators of Spatial Association (LISA) for Empirical Bayes smoothed incidence rates of Oropouche virus in Northeast Brazil. Top panel: Cluster map showing spatial autocorrelation patterns. Bottom panel: LISA significance map indicating municipalities with statistically significant local spatial autocorrelation at $p < 0.05$, 0.01, and 0.001.

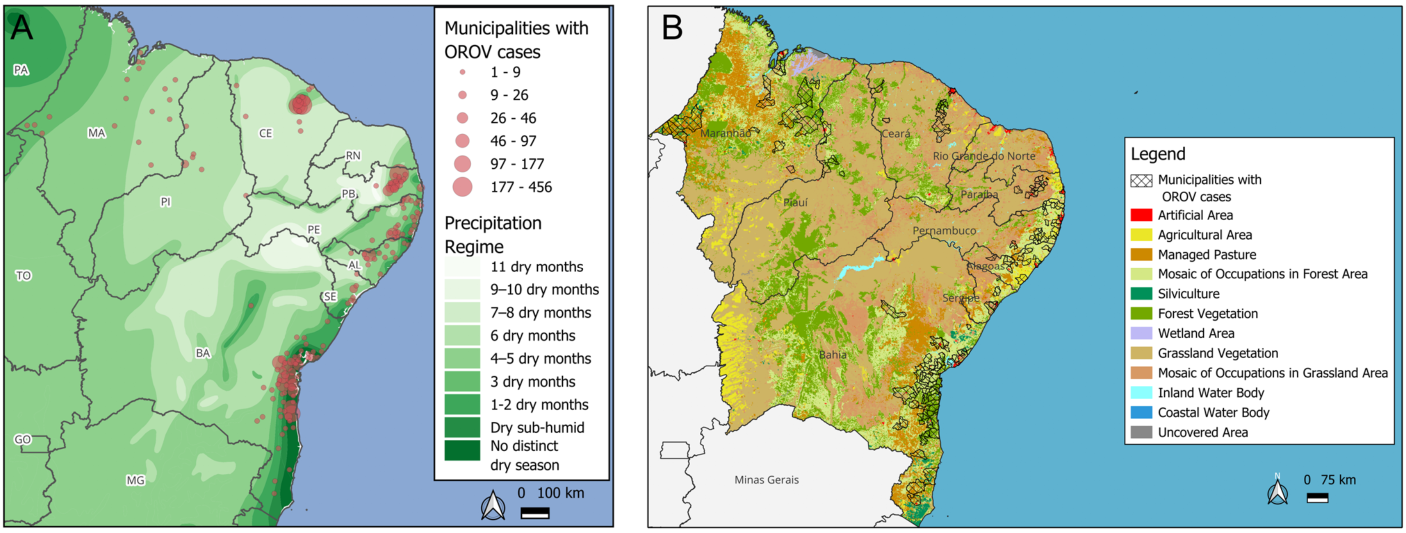


**Figure S5 -** Distribution of municipalities with OROV cases across precipitation regime zones (A) and land cover and use (B) in Northeast Brazil.

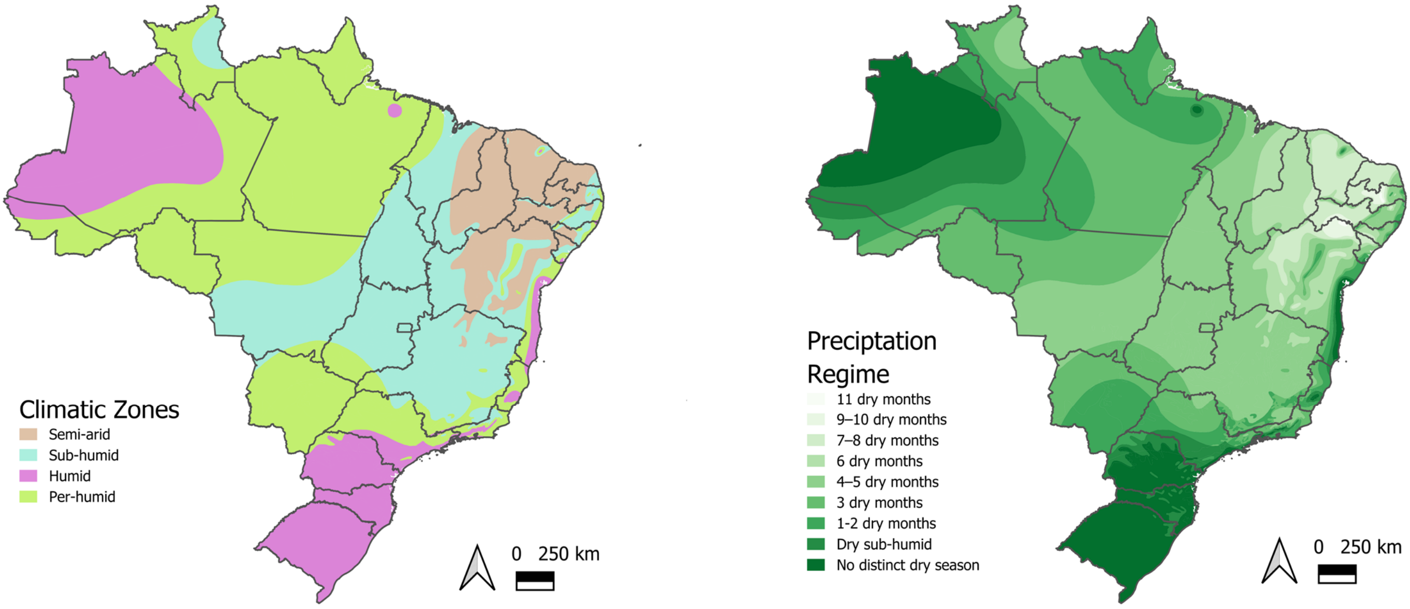


**Figure S6 -** Climatic zones and precipitation regimes across Brazil.

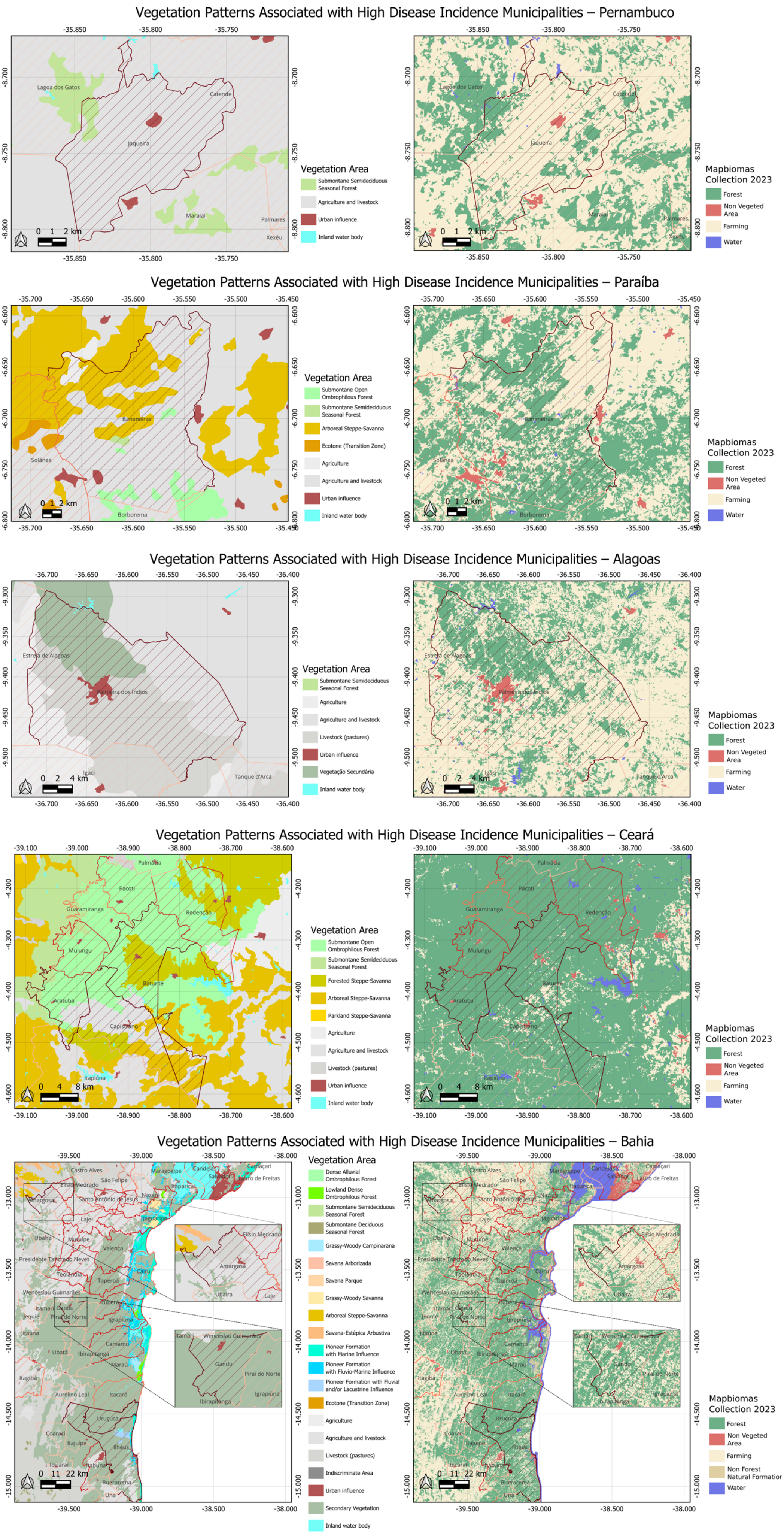


**Figure S7** - Vegetation characteristics of municipalities with high incidence of Oropouche virus in five northeastern Brazilian states. Each panel compares vegetation cover (left) and land use classification from MapBiomas 2023 (right), highlighting the presence of forest fragments, transitional zones, and agricultural areas within affected municipalities.

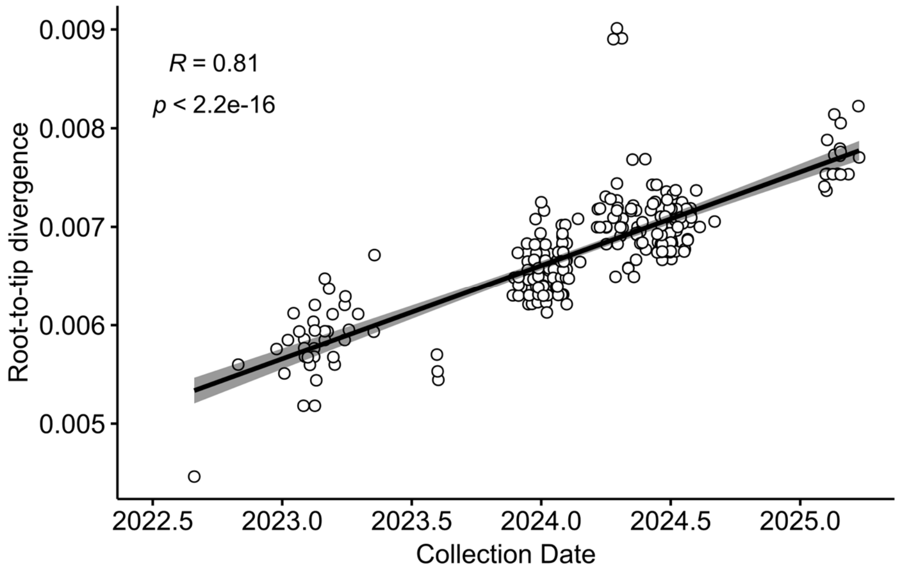


**Figure S8.** Root-to-tip divergence regression of OROV genomes showing a moderate temporal signal (R = 0.81)
